# Plasma Proteomics in Acute Heart Failure: Survival-Associated Protein Signatures and 90-Day Trajectories

**DOI:** 10.64898/2025.12.01.25341310

**Authors:** Alexandra M Chitroceanu, Pauline Fahjen, Daniel Schulze, Veronika Zach, Lina Alasfar, Matthias Ziehm, Marieluise Kirchner, Sylvia Niquet, Jan-Niklas Dahmen, Carlos Plappert, Lucie Kretzler, Andreas Kind, Daniela Zurkan, Rafaela Maria Pinto, Edwardo Reynolds, Tobias D Trippel, Sofia Forslund, Ulrike Grittner, Knut Mai, Michael Gotthardt, Gabriele G Schiattarella, Ulrich Kintscher, Michael Ahmadi, Leif-Hendrik Boldt, Nikolaus Buchmann, Kai-Uwe Eckardt, Matthias Endres, Holger Gerhardt, Norbert Hübner, Jil Kollmus-Heege, Ulf Landmesser, Dominik N Müller, Christian H Nolte, Sophie K Piper, Simrit Rattan, Ira Rohrpasser-Napierkowski, Katharina Schönrath, Jeanette Schulz-Menger, Oliver Schweizerhof, Tobias Pischon, Burkert Pieske, Joachim Weber, Philipp Mertins, Frank Edelmann

## Abstract

**Background:** Acute heart failure (AHF) patients face poor outcomes, particularly within the high-risk 90-day post-discharge phase. Additional biomarkers reflecting the multifaced pathophysiology of AHF are necessary to improve outcome prediction.

**Objective:** This study aimed to identify novel plasma protein biomarkers and biological pathways related to survival and recovery following an AHF event.

**Methods:** Plasma samples were obtained from patients enrolled in the BeLOVE (Berlin Long-Term Observation of Vascular Events) cohort during an AHF event, at 90-day follow-up, and from a reference group (patients with cardiovascular risk but no recent AHF). Proteomics analysis was performed using both Mass Spectrometry (MS) and Olink Explore (Uppsala, Sweden). Cox proportional-hazards regression identified plasma proteins linked to 90-day all-cause mortality post-AHF.

**Results:** Out of 2,324 proteins analyzed from Olink and 533 from MS, 67 were significantly associated with 90-day mortality (|lnHR|>0.4, FDR<0.05). Inflammation, apoptosis, and extracellular matrix remodeling were key mortality predictors, while metabolic proteins, coagulation control, and complement system associated with survival. TNF receptor family members showed promise for risk stratification beyond natriuretic peptides. Comparison of plasma from survivors at acute setting and 90-day follow-up identified 591 significantly altered proteins via Olink and 131 via MS (logFC>75th percentile, FDR<0.05). CA125 (logFC = -1.8 [-2.2,-1.4], FDR ≈ 2×10⁻⁸) emerged as a biomarker for clinical recovery, declining more sharply than NT-proBNP.

**Conclusions:** Combining targeted and untargeted proteomic approaches identifies novel pathways and biomarkers to refine risk stratification in AHF. Profiling of proteomic changes post-AHF provides critical insights into the molecular processes underlying disease progression and recovery.

## Introduction

Acute heart failure (AHF) is a shared clinical presentation of diverse cardiac conditions, with patients exhibiting symptoms and signs of heart failure (HF) with varying degrees of severity.^1^ Although AHF results in similar clinical manifestations, the underlying cardiac conditions and contributing factors can differ significantly leading to highly heterogeneous pathophysiology.^2^ Symptoms, primarily caused by fluid overload and congestion, are managed with decongestive medications tailored to the patient’s hemodynamic status.^1,3^ Despite advancements in treatment, including guideline-directed medical therapy, AHF remains associated with poor outcomes, leaving patients at significant risk of disease progression and adverse events.^4^ This is particularly critical during the 90-day post-discharge period, often referred to as the vulnerable phase, characterized by a spike in event rates and a high risk of poor clinical outcomes.^5^ Within this period, 30% of HF patients are readmitted, and 15% do not survive.^5^ Persistent hemodynamic abnormalities do not fully explain the ongoing progression of HF,^6^ underscoring the need to discover and understand the modifiable molecular pathways driving disease.

Historically, biomarkers such as natriuretic peptides and troponin have been used as indicators of adverse outcomes in HF.^1,4,7^ Recent evidence highlights the need for additional biomarkers,^8^ that reflect the diverse underlying pathophysiology, potentially improving outcome prediction when combined with clinical assessments. Advances in technology now allow the analysis of thousands of circulating proteins through aptamer or antibody-based approaches,^8^ enabling novel biomarker discovery. However, these methods can introduce bias, as they focus on manually preselected proteins included in manually curated panels. To overcome this limitation, integrating deep targeted proteomic phenotyping with untargeted mass spectrometry (MS) provides an unbiased, comprehensive view of the biological state. Coupled with robust clinical data analysis, this integrated approach may provide valuable insights into the determinants of AHF outcomes, progression and recovery, paving the way for personalized and mechanism-based treatments.

## Methods

### Study Cohort

Patients diagnosed with AHF were enrolled in the Berlin Long-Term Observation of Vascular Events Study (BeLOVE; German Clinical Trials Registry DRKS00016852), an ongoing observational prospective clinical cohort study that aims to improve prediction and mechanistic understanding of cardiovascular (CV) disease progression, by comprehensively investigating a high-risk patient population with different organ manifestations. The study was approved by the Charité–Universitätsmedizin Berlin ethics committee (EA1/066/17) and conducted in accordance with the Declaration of Helsinki, with all participants providing informed consent. Recruitment included patients hospitalized for acute CV events or individuals at high CV risk. Initial examinations occurred during the acute phase (day 0-7) of the index CV event or after inclusion into the chronic high-risk subcohort, followed by in-depth clinical phenotyping at approximately 90-days (median = 99d, IQR = 90-112, for included AHF patients). Deep clinical phenotyping included assessments of the patient’s medical history, clinical and laboratory parameters and patient-related outcome measures. Inclusion criteria for the BeLOVE AHF subcohort included hospitalization for AHF with a New York Heart Association (NYHA) class ≥ II or clinical deterioration of chronic HF, along with either the escalation of a pre-existing loop diuretic therapy or a new prescription of loop diuretics. Exclusion criteria comprised reduced life expectancy (<6 months) due to a non-CV cause, active cancer, history of organ transplantation, primary diagnosis of acute coronary syndrome, acute stroke, or acute kidney injury. A different subcohort consists of patients with very high-risk chronic CV conditions who have not experienced an event very recently. For detailed inclusion and exclusion criteria, as well as the study design, refer to the published BeLOVE study protocol.^9^ For this study, we selected 90 patients from the AHF arm of the BeLOVE study based on available biosamples, prioritizing those who died within the observation period and those who survived at least 90 days to provide a second plasma sample at follow-up. Our reference group consisted of 30 patients with very high-risk chronic CV conditions but no AHF event in the past year, recruited between 2018 and 2022. Cardinality matching was used to pair these 30 patients with AHF survivors who had a second plasma sample, matching based on age, sex, and estimated glomerular filtration rate. One reference patient withdrew consent after study inclusion and was removed from all analyses. Study data were collected and managed using REDCap electronic data capture tools hosted at Berlin Institute of Health, Berlin, Germany.^10^ Patient characteristics were exported from the BeLOVE database on May 28, 2024.

Ethylene-diamine-tetraacetic acid (EDTA) plasma samples for proteomic analysis were collected from peripheral venous blood during the acute visit, within 7 days after index event, for all patients with HF, as well as for the reference group. Additionally, samples were collected during the 90-day deep-phenotyping visit for 52 patients with HF.

### Olink targeted proteomic analysis

3072 human plasma proteins were targeted for measurement in EDTA plasma using Olink Explore technology (Olink, Uppsala, Sweden). The selected biomarkers encompassed low-abundant inflammation proteins, proteins actively secreted into the bloodstream, approved and potential drug targets, organ-specific proteins indicative of leakage into circulation, and exploratory biomarkers. The Olink technique employs Proximity Extension Assay technology, where two antibodies with unique oligonucleotide sequences bind to the protein of interest in the plasma sample. The hybridized oligonucleotides are extended into a protein-specific deoxyribonucleic acid (DNA) barcode by DNA polymerase. Subsequently, polymerase chain reaction amplifies the barcode, and its quantity reflects the original protein concentration. Next-generation sequencing is used to read the DNA barcode count, which is translated into Normalized protein expression (NPX) values—a relative log2 transformed expression measure.

### Untargeted Mass spectrometry-based proteomic analysis

Sample preparation was largely automated using an Agilent BRAVO system. Plasma samples were diluted 1:10 with H_2_O, mixed with lysis buffer (2% Sodium deoxycholate (SDC), 20 mM dithiothreitol, 80 mM chloroacetamide, 22 mM Tris, pH 8), and heated at 95°C for 10 minutes. Proteins were digested overnight at 37°C with trypsin and LysC in 50 mM Hepes (pH 8). Acidification with 10% trifluoroacetic acid precipitated SDC, followed by dilution with 1% formic acid (FA) and centrifugation at 1200g for 10 minutes. The supernatant was desalted on C18 cartridges, eluted in 50% acetonitrile (ACN)/0.1% FA, dried, and stored at -20°C.

For analysis, peptides were reconstituted in 3% ACN/0.1% FA, and 1 µg was injected into a Vanquish Neo high-performance liquid chromatography coupled with an Orbitrap Exploris^TM^ 480 mass spectrometer. Peptides were separated on a 20 cm C18 column using a 32-minute gradient at 250 nL/min. MS data were acquired in data-independent acquisition (DIA) mode with 120K resolution for full scans (350–1650 m/z) and 30K resolution for 40 MS2 scans (variable windows, stepped collision energy: 26, 29, 32). The cycle time was 3 seconds.

Peptide identification was performed in Spectronaut (v17.4) using directDIA and the 2022-03 Uniprot database. Carbamidomethyl (C) was set as a fixed modification, with Acetyl (N-term), Deamidation (NQ), and Oxidation (M) as variable modifications.

### Quality control

Proteins detected in <25% of samples or failing quality filters were excluded from Olink, and <15% from MS analyses. Missing MS values were imputed using a Gaussian distribution (0.3x SD, 1.8x downshift). Two patients from the survivor group were excluded from the analysis as outliers in the Olink Quality Control and PCA plot. They had either <500 average matched counts or a Negative Control median deviation exceeding 5 standard deviations from the assay’s predefined value, triggering a warning status.

### Statistical analysis

All statistical analyses were performed using R (version 4.3.2). For matching the “MatchIt” R package was utilized.^11^ Patient characteristics were analyzed using the gtsummary R-package.^12^ We used Cox proportional-hazards regression models to assess the association between proteomics and 90-day survival of AHF patients. Patients alive at 90 days and those who were lost to follow-up were censored. Hazard ratios (HRs) and p-values were calculated for each protein, adjusted for age and sex, and corrected for multiple testing using the Benjamini-Hochberg method (false discovery rate (FDR) < 0.05). For proteins of interest, survival curves were plotted by stratifying AHF patients into three groups based on relative protein abundance (highest quartile, lowest quartile, and intermediate 50%). Pearson correlations of the ln(HR) of proteins identified in both Olink and MS datasets was calculated and scatter plot was generated to visualize the correlation between the two techniques. To reveal proteomic changes during the recovery process, differential abundance was analysed with the limma package,^13^ adjusting for age and sex, when necessary, with statistical significance determined using a FDR threshold of < 0.05. In order to understand systemic protein changes associated with the recovery from an AHF state which can translate to an improved patient re-stratification we performed paired comparisons between the acute and 90-day phases. An FDR of 0.05 was used as significance cut-off. Pathway analysis via Single-sample Gene Set Enrichment Analysis (ssGSEA)^14^ was conducted using HRs and logarithmic fold change (logFC) values from both MS and Olink data. For clarity, redundant pathways with >50% gene overlap to higher-scoring pathways were excluded. To further explore changes in plasma protein levels during recovery from AHF, we grouped proteins with significant differences based on their trajectories across the acute phase, 90-day recovery, and reference group. Mean NPX values were calculated for each protein during the acute phase and at the 90-day recovery visit in survivors, and in the control group. Protein mean NPX values were normalized to the median within each assay for better comparability. Line plots were generated to show protein trends from the acute phase through recovery, compared to controls.

## Results

### Patient Characteristics

The general characteristics of AHF patients during the acute event hospitalization are presented in *Table 1*. Within the 90-day vulnerable phase, 21 patients died from all causes, categorizing them as "non-survivors", in contrast to the 69 patients who survived this period, identified as "survivors". Non-survivors were older (77 vs. 73 years, p = 0.010), were more often female (52% vs. 29%, p = 0.048), had higher N-terminal pro B-type natriuretic peptide (NT-proBNP) levels (4.887 vs. 1.933 pg/ml, p = 0.001), and lower hemoglobin (10.9 vs. 12.5 g/dl, p = 0.007) and albumin levels (35.5 vs. 39.6 g/L, p = 0.004) compared to survivors. Other characteristics showed no substantial differences.

In survivors, improvements were observed from the acute phase to 90 days. NYHA class improved (67% of patients improved, p < 0.001) and NT-proBNP levels decreased (1.931 to 1.040 pg/ml, p = 0.001). Albumin levels increased (40.3 to 43.5 g/dL, p < 0.001). Reductions in alamin transaminase (ALT), gamma-glutamiltransferase (GGT), lactate dehydrogenase (LDH), bilirubin, ferritin and sodium were also noted, while potassium and cholesterol levels increased.

The reference group consisted of individuals at very high CV risk, without an AHF event in the last 12 months (see Methods – Study cohort). The reference group was matched based on age (74 years (61-80), vs. 73 years (63-87), p = 0.8) and sex (31% female vs. 29% female, p > 0.9) with the AHF survivor group. *Supplementary tables 1 and 2* provide detailed comparisons, also with the reference group.

### Plasma proteomic predictors of short-term outcomes following an AHF event

Out of the 3072 proteins targeted, quantification was successfully achieved for 2901 proteins. Following filtering to remove proteins failing Olink quality control, those with duplicated measurements, or NPX values below the protein-specific limit of detection in over 25% of samples, 2324 unique proteins remained for further statistical analysis. 533 proteins were identified through the untargeted MS approach, including 308 that are not covered by the Olink panel. Cox proportional-hazards regression models identified 65 plasma proteins associated with 90-day mortality after an AHF event using the Olink targeted approach. Among these, 58 exhibited HR greater than 1.5 (HR range: 1.6 - 6.5, FDR < 0.05), indicating a positive relationship where higher relative protein concentrations were associated with an increased mortality rate, while 7 proteins showed significantly low HR (HR range: 0.07 - 0.4, FDR < 0.05), and therefore an association with survival (*Figure 1.A*). Additionally, the untargeted MS approach uncovered two more associations, ITIH3 (Inter-alpha-trypsin inhibitor heavy chain H3) and LMAN2 (Lectin, Mannose Binding 2). The top proteins positively associated with mortality risk were ITIH3, LMAN2, B4GALT1 (Beta-1,4-galactosyltransferase 1), MCFD2 (Multiple coagulation factor deficiency protein 2), and IGFBP4 (Insulin-like growth factor-binding protein 4). Conversely, the top proteins linked to lower mortality risk included BTD (Biotinidase), AFM (Afamin), INHBC (Inhibin beta C chain), F12 (Coagulation factor XII), and SERPINA4 (Kallistatin). A list of the 20 most strongly associated proteins identified as potential outcome predictors and their potential role in CV disease in *Supplementary Table 3*. The Pearson correlation of ln(HR) for overlapping proteins between Olink and MS is 0.66, confirming a moderate agreement between the two techniques and validating our markers. Notably, ITIH3 is the only protein identified as significant in the MS-based regression analyses that was not validated by Olink analysis (*Supplementary Figure 1*). Kaplan-Meier survival curves were used to compare protein levels and mortality in the 90 days post-AHF *(Figure 1.B)*. Higher NT-proBNP levels (top panel) displayed only a trend toward lower survival rates (lnHR = 0.6 [0.1, 1.1], FDR = 0.11) and showed less discrimination ability (C-index 0.74) among the protein levels than other identified markers (*Supplementary Table 3*). For TNFRSF10A (TNF receptor superfamily member 10a - bottom panel), the group separation is clearer, particularly in the first 90 days, with patients in the highest quartile showing a markedly lower survival probability, surpassing the predictive value of NT-proBNP for early mortality (C-index 0.78).

**Figure 1:**
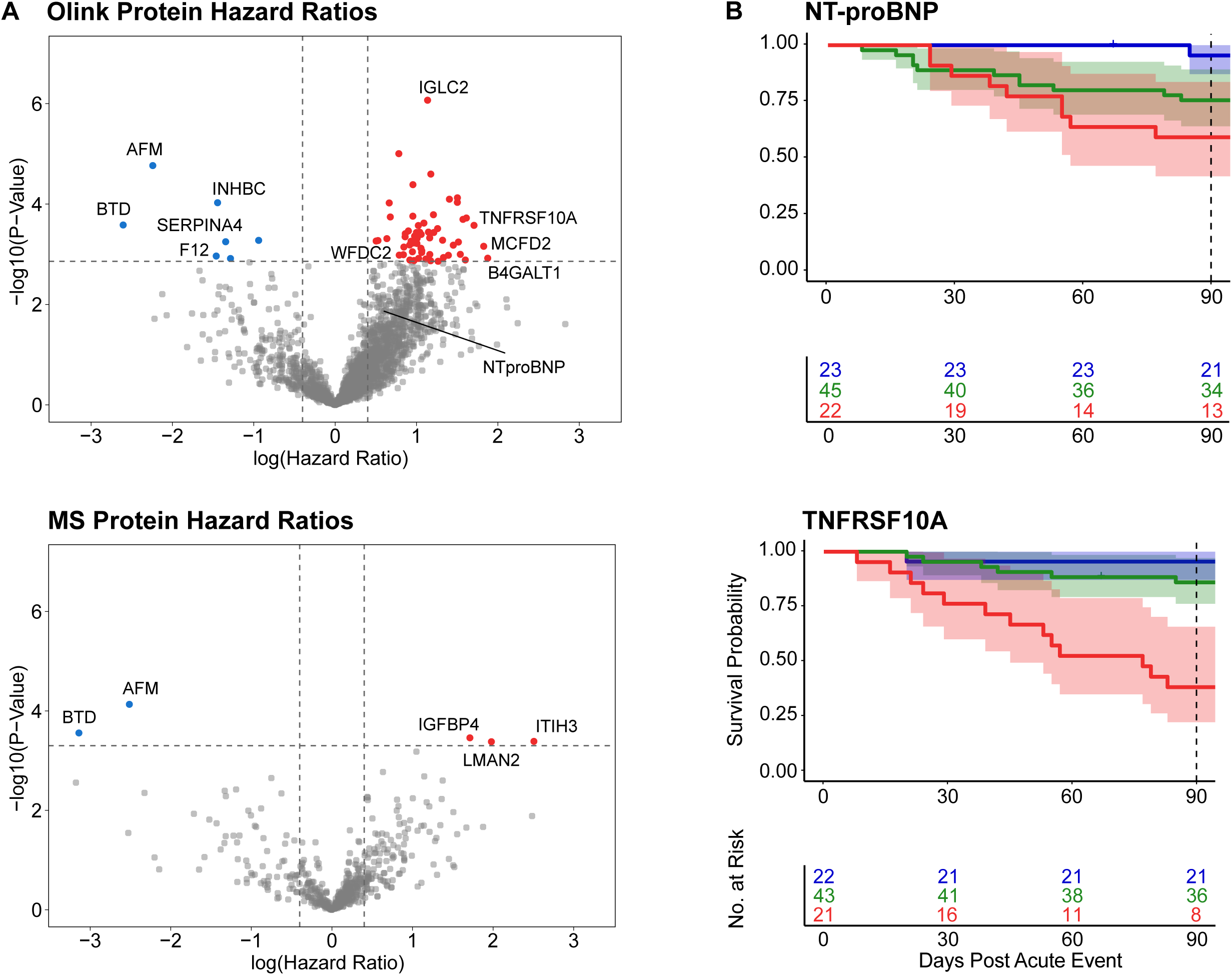
Survival Analysis. **A.** Volcano plots showing plasma protein associations with 90-day mortality post-AHF, measured via Olink (top) and MS (bottom). Age- and sex-adjusted log hazard ratios are displayed, with FDR < 0.05 as significant. Blue proteins indicate improved survival, red indicate increased mortality risk. **B.** Kaplan-Meier 90-day survival curves for NT-proBNP and TNFRSF10A, with patients stratified into quantiles by Olink protein level at hospitalization: top 25% (red), middle 50% (green), and bottom 25% (blue) with 95% Confidence intervals. The curves highlight the prognostic value of these markers for AHF outcomes. Differences in patient numbers reflect variations in data availability for Olink measurements. Novel identified markers surpass the predictive power of NT-proBNP in the analyzed cohort. *AHF: Acute heart failure. FDR: False discovery rate. MS: Mass spectrometry. NT-proBNP: N-terminal pro B-type natriuretic peptide*.

The ssGSEA analysis indicated that proteins associated with a higher probability of mortality were predominantly linked to systemic inflammation, immune response, extracellular matrix (ECM) remodeling, and apoptosis. In contrast, proteins associated with a lower risk of death were primarily involved in metabolic processes, the complement system and coagulation (*Figure 2)*.

**Figure 2:**
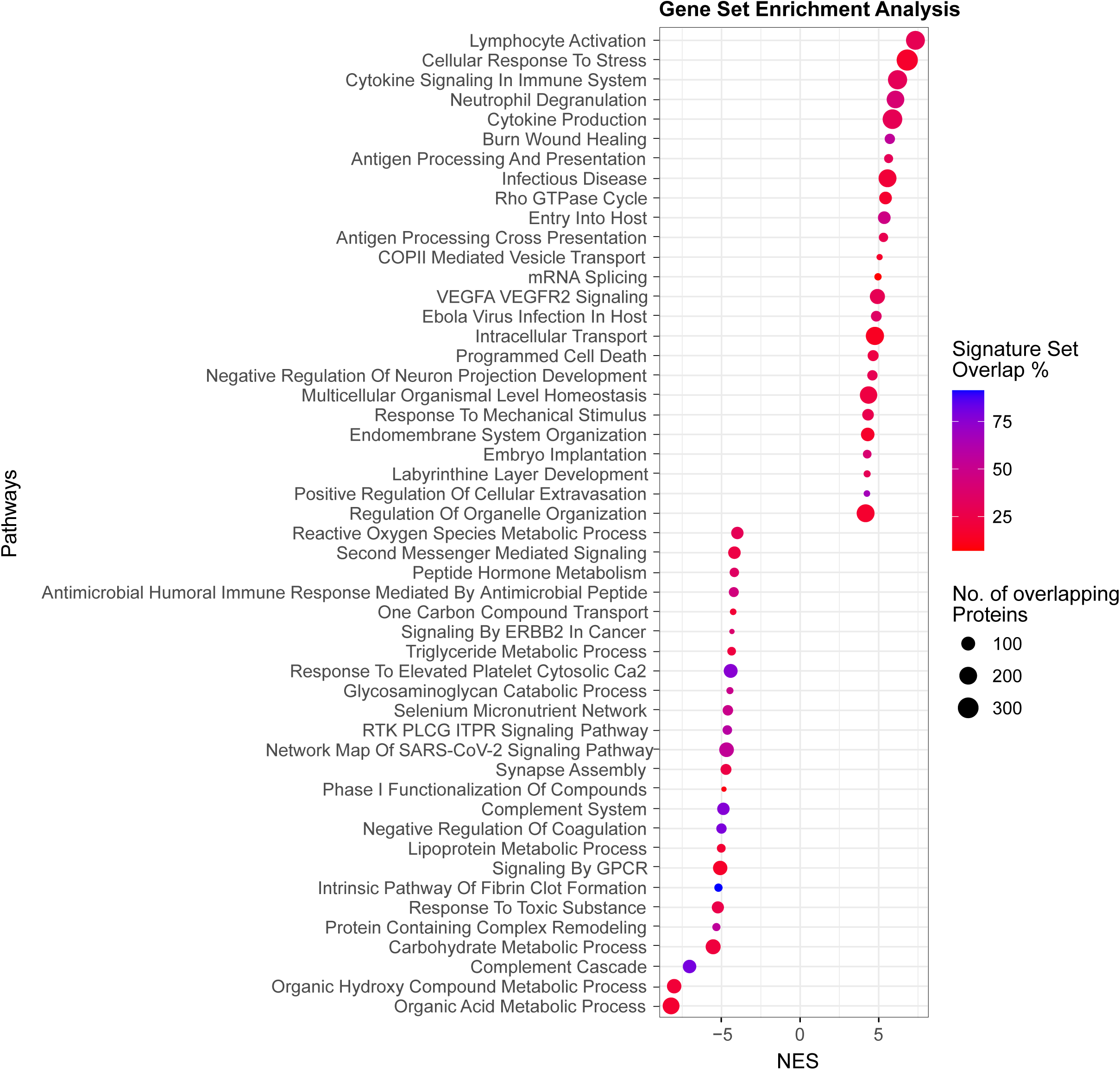
Biological Pathways related to 90-day Outcome. Dot plot of ssGSEA results from combined MS and Olink hazard ratios, highlighting the top 25 non-redundant significant (FDR < 0.05) pathways linked to survival and mortality. The x-axis shows NES, with negative values indicating higher activation of pathways associated to lower mortality rates and positive values to higher mortality rates. Dot size reflects the number of observed proteins in the gene set, while colour intensity represents the percentage of the gene set’s proteins that were observed. Inflammation and immune activation and reduced levels of circulating metabolic proteins are associated with worse outcomes. *FDR: False discovery rate. MS: Mass spectrometry. NES: normalized enrichment scores. ssGSEA: single sample gene set enrichment analysis*.

### Plasma proteome dynamics during the recovery phase after AHF

To gain a deeper understanding of systemic molecular changes during the recovery phase, we compared plasma proteome profiles of the survivor group during the acute event and at the 90-day follow-up (*Figure 3.A*). The Olink analysis identified 591 proteins with significantly different abundance, with absolute logFC ranging from 0,3 – 1.8. The MS analysis resulted in 131 significantly changing proteins (|logFC| range: 0.2 – 1.6, FDR < 0.05), of which 11 were also significant in the Olink data and 118 more were quantified but did not pass the significance cut-off in Olink.

**Figure 3:**
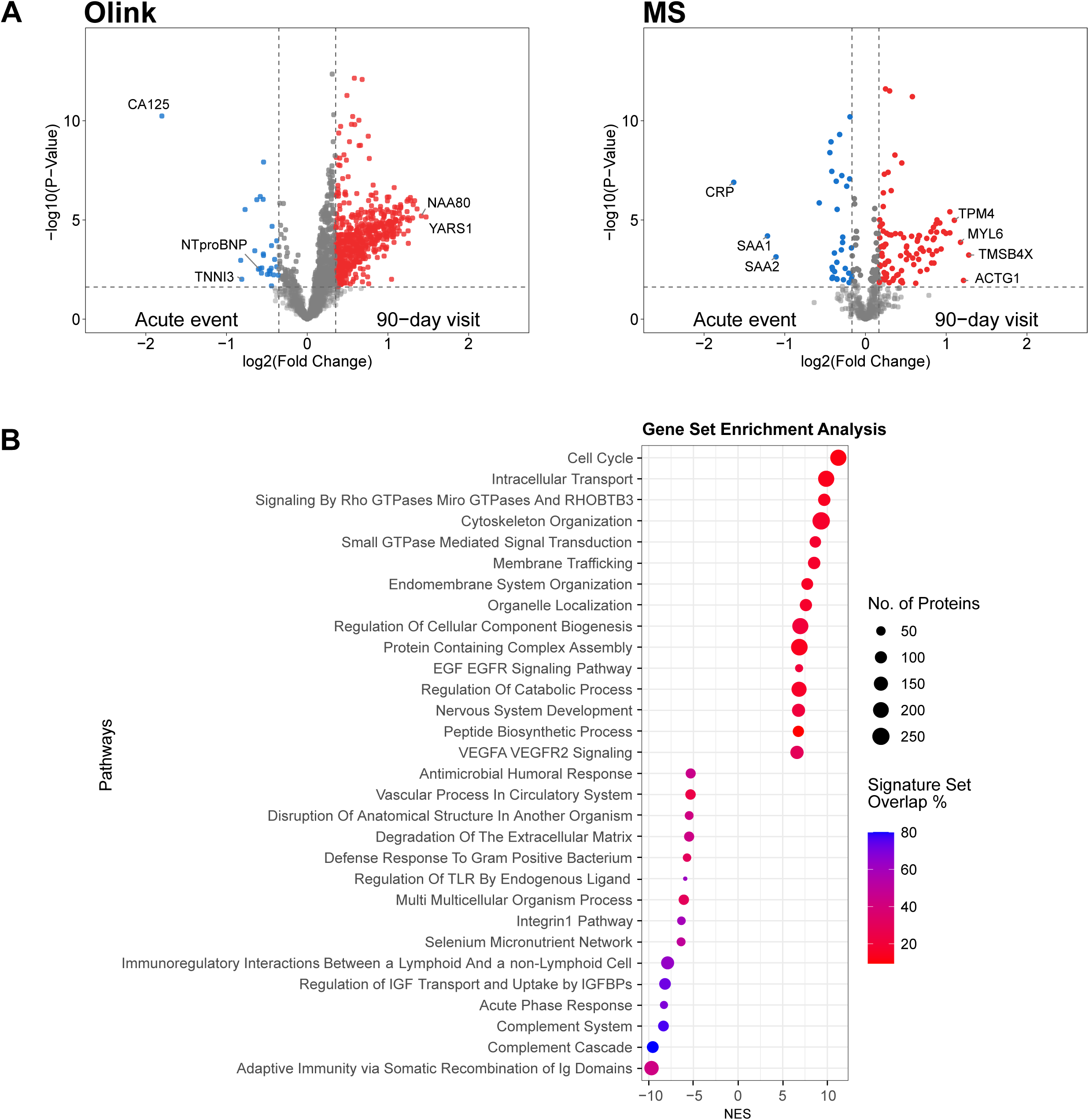
Plasma Proteomic Differences between Acute and Recovery Phase. A. Volcano plots showing differential protein abundance between acute hospitalization and 90-day recovery in survivors with two visits (Olink left, MS right). Paired analysis (n = 52), with logFC. Proteins with FDR < 0.05 are significant and shown in colour. Blue: elevated during the acute phase, red: elevated after 90 days. Congestion marker CA125 displays the strongest decline within the 90 days post AHF discharge. B. ssGSEA of combined MS and Olink logFC showing biological pathway changes between acute and 90-day recovery. The x-axis represents NES, with positive values indicating 90-day enrichment and negative values indicating acute-phase enrichment. Dot size reflects the number of observed proteins, and colour intensity indicates the percentage of observed proteins in the gene set. *AHF: Acute heart failure. FDR: False discovery rate. logFC: logarithmic fold change. MS: Mass spectrometry. NES: normalized enrichment scores*.

Proteins with the greatest reductions after 90 days included CA125 (Cancer antigen 125) also known as MUC16 (Mucin-16), CRP (C-reactive protein), SAA1 (Serum amyloid A-1 protein), SAA2 (Serum amyloid A-2 protein), NPPB (Natriuretic peptides B), and TNNI3 (Troponin I, cardiac muscle), with CA125 and CRP exhibiting the most pronounced decrease in the acute phase, by some distance.

Proteins with higher abundance after 90 days separated into two groups: one with statistically robust but modest changes and another with larger changes but greater variability. The elevations with the lowest FDR were observed in CRTAC1(Cartilage acidic protein 1), CA6 (Carbonic anhydrase 6), HMCN2 (Hemicentin-2), APOA1 (Apolipoprotein A 1), and GSN (Gelsolin). In contrast, proteins with the largest relative elevations included YARS1 (Tyrosine--tRNA ligase, cytoplasmic), NAA80 (N-alpha-acetyltransferase 80), PRKG1 (cGMP-dependent protein kinase 1), APPL2 (DCC-interacting protein 13-beta), and RAB11FIP3 (Rab11 family-interacting protein 3).

Pathway analysis revealed that following an acute event, the transition to a compensated chronic HF state involves a shift from an initial inflammatory and neurohormonal response to processes focused on cellular homeostasis, metabolic and vascular repair *(Figure 3.B*).

### Proteomic shifts toward an improved, compensated state during the 90-day recovery phase after AHF

Proteomic analysis comparing the reference group with survivors at both the acute phase and after 90 days demonstrated a pattern of protein expression changes. During the acute phase, 67 proteins were significantly altered compared to the reference group, with absolute logFC ranging from 0.2 to 2.3, based on combined MS and Olink data. After 90 days, although similar fold changes were observed, these differences were no longer statistically significant. This suggests that the proteomic profile of survivors gradually shifts toward a more compensated state over time, with higher variability in the plasma proteome, reflecting recovery dynamics (*Supplementary Figure 2*).

To further understand the impact of the recovery process on the plasma proteome, proteins that exhibited substantial, significant changes in the comparison acute to recovery phase were categorized into eight distinct profiles based on their mean level trajectory from the acute phase to the 90-day follow-up, relative to the reference group (*Figure 4*). Profiles 1-4 highlight proteins that significantly decreased during the recovery phase in the surviving patients. Profile 1 includes five proteins, such as CA125 and NT-proBNP, that partially recovered by the 90-day follow-up but did not reach levels observed in the reference group. Profile 2 contains 15 proteins, including acute-phase proteins (CRP, SAA1, SAA2) and muscle-derived proteins (TNNI3, MYLPF, MYBPC1), which decreased to levels below the reference group average. Profile 3 consists of four proteins (APCS, LBP, MYBPC2, PTS) that decreased during recovery, despite being higher in the reference group. Profile 4 features 35 proteins, including acute-phase proteins (SERPINA1, SERPINA3, ORM1), that returned to levels comparable to the reference group.

**Figure 4:**
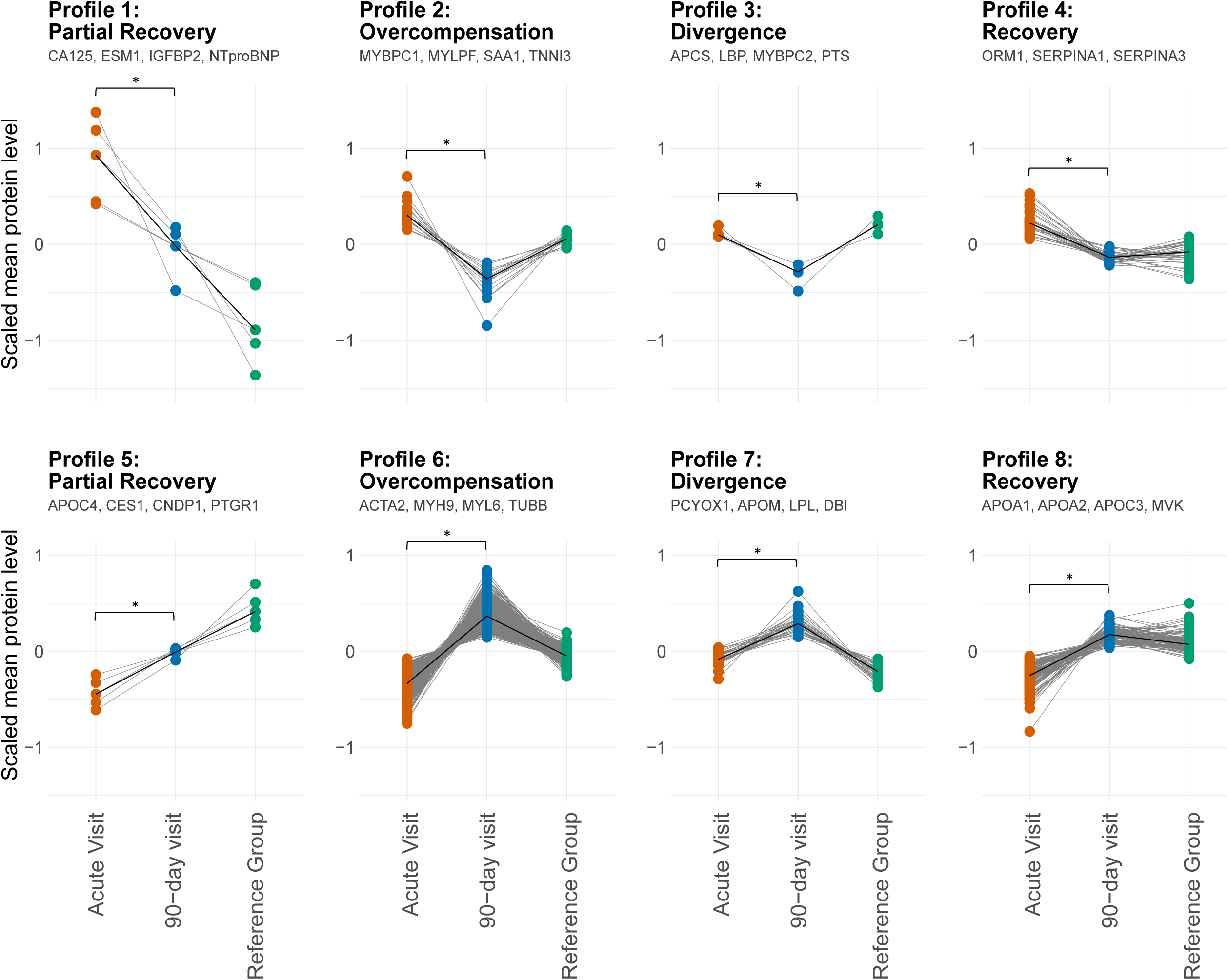
Plasma Proteomic Dynamics in 90-day Recovery. Trends of plasma proteins showing significant changes in the paired analysis between the acute phase and 90-day recovery in survivors, based on combined Olink and MS data. Proteins are categorized into eight profiles based on their normalized abundance changes during the 90-day recovery phase, relative to the reference. For each profile, a subset of proteins is shown, exemplifying the profile. Grey lines represent individual protein trajectories, while black lines denote the median trend for each profile. *MS: Mass spectrometry*.

In contrast, profiles 5-8 represent proteins that significantly increased during recovery. Profile 5 includes five proteins (APOC4, CES1, CNDP1, PTGR1, SSC4D) that showed partial normalization, moving closer to but not fully aligning with reference group levels. Most proteins followed the pattern seen in profile 6, with levels exceeding the reference group, and were mainly involved in cytoskeleton organization (ACTA2, ACTB, ACTBL2, ACTN1, CAP1, CFL1, MYH9, MYL6, MYL9, TUBA1B, TUBA4A, TUBB, TUBB1, VCL, TLN1, WASF1, GIT1, SPARC).

Profile 7 comprises 34 proteins, including four linked to lipid metabolism (PCYOX1, APOM, LPL, DBI) that increased during recovery even though they showed lower levels in the reference group. Profile 8 highlights 142 proteins, mainly related to metabolic processes (ALDH5A1, APOA1, APOA2, APOA4, APOC1, APOC3, GNAS, GSTA1, GSTO1, MVK, NIT2, PGK1, PKM, PON1) that returned to the reference group levels.

## Discussion

Timely risk stratification in AHF during hospitalization is crucial for guiding clinical interventions and improving post-discharge outcomes. Our study analyses systemic plasma proteomic changes during the recovery phase, from the acute event to 90 days post-discharge, with two key objectives: 1) identifying proteins associated with survival based on their acute-phase levels and 2) exploring plasma proteomic changes 90 days after an AHF event to discern pathways of AHF improvement. This is the first comprehensive evaluation of time-dependent protein alterations in AHF patients, offering significant insights into progression and recovery mechanisms. Our findings are supported by the identification of proteins previously linked to CV disease prediction or HF-related pathophysiology and are also technically validated by a good correlation between two different proteomic techniques. Additionally, we identified proteins not previously associated with these conditions, with unknown functions, as shown in *Supplementary Table 3*.

### Plasma proteomic predictors of short-term outcomes following an AHF event

Proteins related to **persistent systemic inflammation and immune processes** are the main factors influencing the survival status in our regression analysis, which in turn accelerate **cell apoptosis** and **necrosis processes**. Elevated levels of proinflammatory cytokines are typically correlated with disease severity and poorer prognosis in AHF patients.^15^ In non-survivors, we observed stronger proinflammatory signaling, and an activation of lymphocytes, particularly B-cells, and neutrophils, which are known to release cytokines contributing to cardiomyocyte apoptosis and cardiac remodeling.^16^

Among the proteins that have previously been connected to HF outcome, several members of the tumor necrosis factor (TNF) receptor family (TNFRSF1B, TNFRSF13B, TNFRSF17, TNFRSF10A and TNFRSF10B) were linked to survival outcomes in this study. TNF receptors are cell surface proteins involved in apoptotic, necroptotic, and inflammatory signaling pathways.^17^ Notably, TNFRSF10A acts as a receptor for TNF-related apoptosis-inducing ligand (TRAIL) in death receptor signaling.^18^ Increased concentrations of TNFRSF10B (TNF receptor superfamily member 10B) also known as TRAIL-R2 (TNF-related apoptosis-inducing ligand receptor 2) in particular, have been associated with all-cause mortality and incident HF in previous proteomics HF studies.^19,20^ LILRB2 (Leukocyte immunoglobulin-like receptor subfamily B member 2), is another protein of interest identified in our analysis. It has been implicated in the regulation of immune responses, especially through intracellular signaling pathways^21^ and emerging evidence links it to acute myocardial infarction progression, highlighting its potential as a marker for CV disease.^22^ In addition to established CV biomarkers, novel proteomic markers identified in this study, reflecting inflammation, immune response, and apoptosis, include SLAMF1, SLAMF7, CD4, CD27, CD79B, FCRL5, FCRLB, BATF, BAG4, BNIP3L, BCL2L1, ZBP1, GADD45GIP1, MZB1, and NBN.

Moreover, our analysis revealed that proteins which were associated with worse outcomes, are linked to **impaired ECM remodeling** and subsequent **fibrosis.** Excessive ECM accumulation impairs cardiac contractility and increases the risk of arrhythmia.^23^ Furthermore, ECM remodeling is a key feature of HF, and increased levels of ECM turnover markers have previously been observed in the bloodstream of patients with AHF.^24^ Among the identified proteins linked to a worse 90-day outcome is TXNDC5 (Thioredoxin domain-containing protein 5), which is highly expressed in cardiac fibroblasts and promotes fibrosis by enhancing ECM protein folding. Modulating TXNDC5 could therefore offer a novel therapeutic avenue for reducing fibrosis and improving outcomes in HF patients.^23^ Upregulation of Tenascin-C (TNC), a large glycoprotein re-expressed by the cardiac fibroblasts under pathological conditions, may be linked to detrimental ventricular remodelling through its proinflammatory and profibrotic effects.^25^ Given its specificity to cardiac inflammatory processes, TNC could be a valuable diagnostic biomarker for assessing heart inflammation and fibrosis.^25,26^ Another key protein identified in this study is WFDC2 (WAP Four-Disulfide Core Domain 2), also known as HE4 (Human Epididymis Protein 4), which has been shown to promote cardiac fibrosis.^27^ WFDC2 has emerged as a promising biomarker for HF outcomes, both in chronic and acute settings.^28,29^ Insulin-like growth factor binding proteins (IGFBPs) are widely studied across various pathologies, and some have shown associations with all-cause mortality in HF patients.^30^ However, their biomarker potential is limited due to the complex regulation they undergo, including hormonal control, proteolytic degradation, and posttranslational modifications.^31^ Novel proteomic outcome markers related to ECM remodelling and fibrosis include ADAMTS4, ADAMTS16, ELN, EPPK1 and KLK4.

The pathway analysis also reveals higher levels of metabolic proteins were associated with higher survival rates. However, these associations should be carefully interpreted. Proteins associated with survival might simply be lower in non-survivors, not necessarily providing protective effects. To better understand this, comparing protein levels with those in a non-acute HF state is essential. In our study, proteins with significantly negative HRs were consistently found at higher levels in the reference group compared to AHF patients (*Supplementary Figure 2*). This suggests that these proteins reflect biological processes that are diminished in non-survivor patients with HF, rather than serving as protective mechanisms. However, it remains unclear whether the observed effects are causal of death or a consequence of the end-stage disease phenotype, potentially driven by systemic dysfunction, including liver impairment.

Additional pathways highlight the critical role of regulating **coagulation processes** to maintain homeostatic balance in HF. Effective coagulation control is not only crucial for ensuring homeostasis, preventing excessive bleeding and clotting issues but also significantly impacts patient survival outcomes. Interestingly, we also observed a higher abundance of **complement system** proteins in patients with better outcomes, potentially reflecting their role in myocardial repair.^32^ Complement proteins such as F12 have been also connected to processing and release of bradykinin which preserves CV function, arterial vasodilation and improved LV relaxation.^33,34^

### Plasma Proteomic dynamics during the recovery phase after AHF

To gain a deeper understanding of the protein dynamics involved in HF improvement, we analysed proteome changes in HF survivors in the acute phase and after a 90-day recovery period and compared these with a reference group of patients at high CV risk but without a recent acute event. Our findings demonstrate significant proteomic changes during recovery, reflecting overall patient improvement. By 90 days, plasma protein levels in AHF survivors show no significant differences from the reference group, contrasting sharply with the pronounced differences observed between the acute phase and the 90-day follow-up.

Three predominant mechanisms characterized the **acute HF phase:** heightened **immune-inflammatory responses**, **myocardial injury** and **necrosis,** and **congestion** with **neurohormonal activation.** These processes were indicated by elevated levels of acute-phase proteins such as CRP, troponins, and natriuretic peptides, which are well-established biomarkers in HF.^35^ Emerging evidence also points to CA125^36^ and serum amyloids^37^ as potential novel inflammatory markers in HF. Our analysis supports this, revealing that both CA125 and SAA levels were significantly elevated during the acute event.

CA125, a glycoprotein synthesized by mesothelial cells, plays a role in cell-mediated immune responses and cardiac remodeling.^36^ Elevated levels of CA125 are associated with congestion in HF, reflecting its potential role in tracking disease progression after decompensation.^36^ Our findings align with the literature, showing a sharp reduction in CA125 levels after 90 days of recovery, a reduction that surpasses even that of NT-proBNP. Both CA125 and NT-proBNP remained elevated compared to the reference group, suggesting that even at 90 days, these patients may still be in a more congested and vulnerable state, as illustrated by the Profile 1 trajectory in *Figure 4*.

SAA is a sensitive marker of acute inflammation, and recent evidence suggests a strong relationship between SAA levels and future CV events in coronary artery disease, similar to CRP.^37^ Additionally, recent findings suggest a potential role of SAA as an inflammatory and fibrotic mediator between the liver and heart in HFpEF, possibly contributing to inflammation driven by metabolic cues.^38^

As patients move through the recovery period, their proteomic profile shifts. Our analysis of survivors shows a transition toward enhanced control of **inflammatory processes, cytoskeletal reorganization**, and **enhanced angiogenesis,** alongside normalization **of metabolic processes**. Proteins associated with cardiac and skeletal muscle damage, such as **TNNI3**, **MYLPF**, **MYBPC1**, **MYL3**, **ACTN2**, and **MYOM3**, showed elevated levels during the acute phase but significantly declined by 90 days, indicating effective tissue healing. Cytoskeletal remodelling is essential for maintaining cellular homeostasis and adapting to mechanical stress and injury in HF.^39^ Our results indicate that 90 days after an AHF event, proteins involved in cytoskeleton organization, such as gelsolin (GSN) and NAA80, are elevated in plasma. Gelsolin regulates actin filament dynamics, and has been shown to be overexpressed in failing human hearts,^39^ while NAA80 modulates actin N-acetylation.^40^ Overexpression of NAA80 at 90 days suggests a potential protective mechanism against protein degradation and cytoskeletal reorganization, supporting enhanced survival.^41^

Conclusions: In summary, our findings show that persistent inflammation, apoptosis, and prolonged ECM remodeling with fibrosis are key predictors of mortality in AHF patients. Pathways connected to metabolism, coagulation, and the complement system are associated with survival. In contrast, enhanced inflammation control, reduced congestion, improved neurohormonal balance, and cytoskeletal reorganization are the main drivers of the recovery process, with CA125 emerging as a key marker for recovery. This study is one of the first to present a detailed proteomic analysis focused on the post-AHF phase, identifying a comprehensive set of novel biomarkers, specific for the vulnerable phase, which warrant further validation in multi-center cohorts. Notably, our results align with current clinical practice and existing literature, where easily detectable biomarkers such as NT-proBNP, CRP, and troponins remain well-established indicators of disease status. In addition, some of the markers we identify including WFDC2 (HE4), a promising marker of HF-related fibrosis, are already measurable through established immunoassays. This reinforces their potential as clinically relevant biomarkers for monitoring AHF progression and recovery, bridging the gap between novel discoveries and existing diagnostic tools. By complementing the targeted Olink approach, MS adds valuable breadth and depth to the proteomic analysis, enhancing the discovery of novel molecular mechanisms and pathways. Incorporating proteomic data into clinical risk stratification algorithms could enable personalized treatments targeting the specific molecular mechanisms driving AHF in individual patients.

## Clinical perspectives

The 90-day follow-up period after an AHF event represents a particularly vulnerable phase characterized by increased risk of adverse outcomes. A comprehensive understanding of the clinical and molecular profiles during this critical interval could facilitate improved risk stratification and guide targeted therapeutic interventions.

## Translational Outlook

Proteomic analysis in AHF identifies 43 key proteins linked to 90-day mortality, emphasizing inflammation, apoptosis, and fibrosis as critical pathways. Metabolic processes, coagulation, and complement activation proteins predict better survival. Novel biomarkers like TNFRSF10A/B enhance risk assessment beyond traditional natriuretic peptides. Based on our findings a multimarker panel that captures the key processes identified will likely be the most effective approach for predicting short-term outcomes in AHF patients. Notably, some of our outcome-associated proteins, including HE4, TNFRSF10B, LILRB2, and TNC, are already easily measurable with available ELISA kits, making clinical translation more feasible. However, validation in an independent cohort is essential, and the absolute levels of these markers in patients must be determined before they can be implemented in clinical practice. Recovery-associated proteomic trajectories indicate reduced inflammation, metabolic restoration, and cytoskeletal reorganization after 90 days, with CA125 emerging as a sensitive marker of clinical improvement. These findings offer novel therapeutic targets and opportunities for personalized patient management.

## Limitations

Our study provides valuable insights into the proteomic dynamics of AHF and identifies novel biomarkers associated with 90-day outcomes; however, several limitations should be acknowledged. The sample size was relatively modest, consisting of 90 AHF patients and 29 reference individuals, which may limit the statistical power and generalizability of the findings. There is also a potential selection bias due to our focus on patients with AHF who did not survive the observation period and those who completed the 90-day follow-up. The single-centre design could introduce an additional selection bias and may not fully represent the broader AHF population. Nevertheless, the integration of targeted and untargeted proteomic approaches and the longitudinal study design of paired acute phase and 90-day follow up visit sampling allowed us to capture a comprehensive proteomic profile during a critical period that is rarely studied, contributing valuable information to the understanding of AHF progression and recovery.

## Data Availability

All data produced in the present study are available upon reasonable request to the authors

## Acknowledgements

The BeLOVE study is substantially funded by the Berlin Institute of Health (BIH at Charité). We thank all participants who took part in the BeLOVE study and the staff in this research program. We would also like to thank the ZeBanC (Zentrale Biomaterialbank der Charité) for the prompt provision of samples and their kind support, and the SFB-1470 and DZHK for funding the entire project. The manuscript was refined in parts for clarity and English grammar using ChatGPT (OpenAI). The final content was reviewed and revised by the authors.

## Abbreviation and acronyms

ACN =: acetonitrile
AHF =: acute heart failure
ALT =: alanine transaminase
BeLOVE =: Berlin Long-Term Observation of Vascular Events
CRP =: C-reactive protein
CV =: cardiovascular
DIA =: data-independent acquisition
DNA =: deoxyribonucleic acid ECM = extracellular matrix
EDTA =: ethylene-diamine-tetraacetic acid
FA =: formic acid
FDR =: false discovery rate
GGT =: gamma-glutamyl transferase
HF =: heart failure
HR =: hazard ratio
LDH =: lactate dehydrogenase
logFC =: logarithmic fold change
MS =: mass spectrometry
NYHA class =: New York Heart Association class
NPX =: normalized protein expression
NT-proBNP =: N-terminal pro B-type natriuretic peptide
PCA =: principal component analysis
ssGSEA =: single-sample gene set enrichment analysis
SD =: standard deviation
SDC =: sodium deoxycholate

---

**Supplementary Figure 1: Correlation of proteins measured by Olink and MS.**

**A** Venn diagram of overlapping proteins quantified using the Olink and MS approach.

**B** Scatterplot showing the ln(HR) of proteins that overlap between the MS and Olink analyses, based on their MS intensities and Olink NPX values. Proteins significantly associated with 90-day outcomes in the Olink analysis are marked in blue, those significant in the MS analysis in red, and proteins identified as significant in both analyses in purple.

*HR: Hazard ratio. MS: Mass spectrometry. NPX: Normalized protein expression*.

**Supplementary Figure 2: Plasma Proteomic Differences between Reference Group and Survivors.**

Volcano plots showing plasma protein differences between the Reference group and Survivors during the acute phase and at 90-day follow-up respectively, measured via Olink (A) and MS (B). log_2_FC are displayed, with FDR < 0.05 as significance cut-off. *FDR: False discovery rate. MS: Mass spectrometry*.

