## Supplementary figures and images for "Plasma Proteomics in Acute Heart Failure: Survival-Associated Protein Signatures and 90-Day Trajectories"

### Supplementary Figure 1

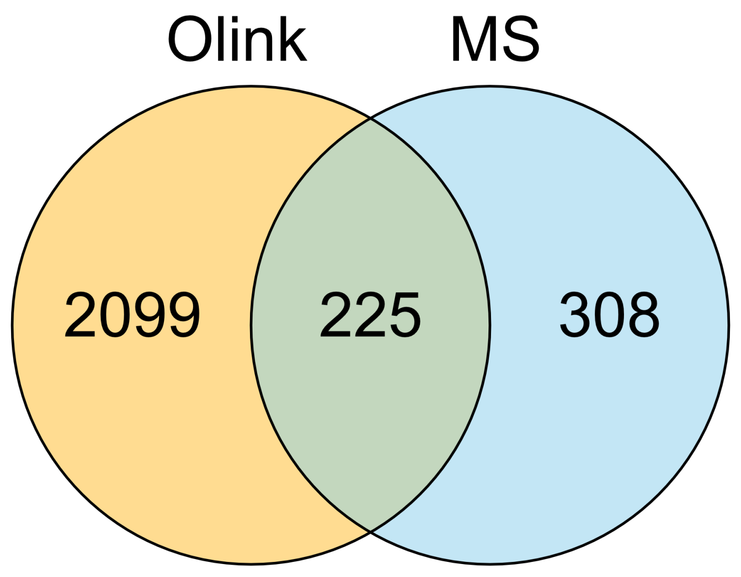

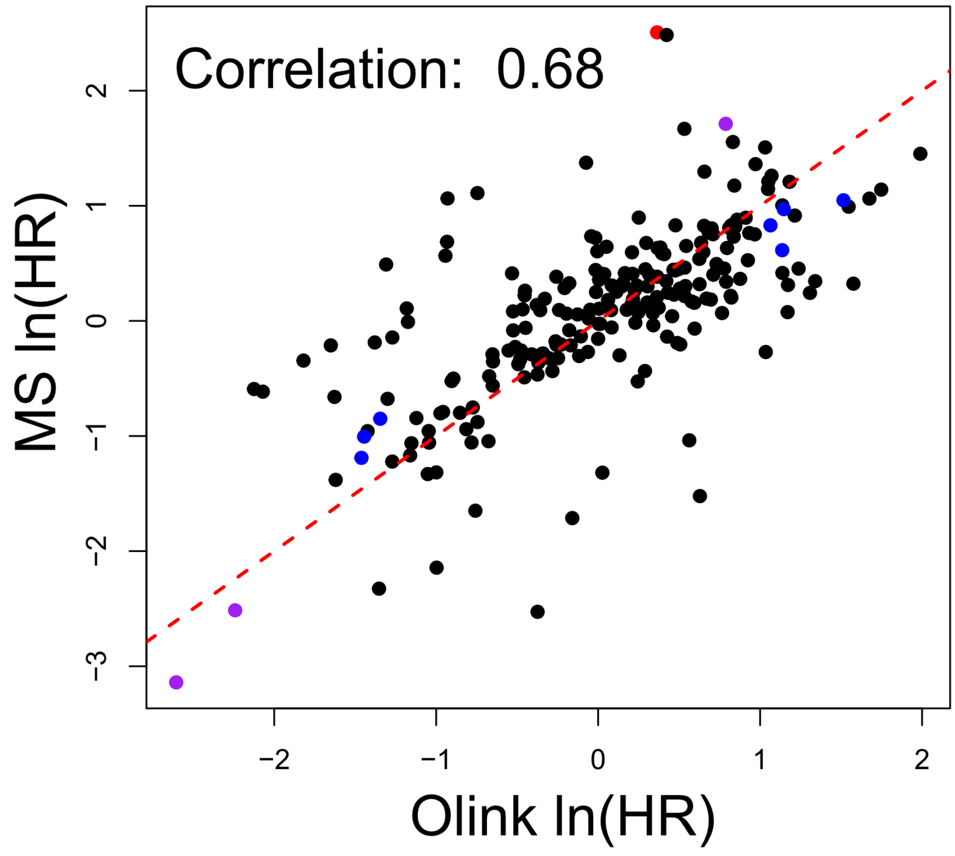

### Supplementary Figure 2

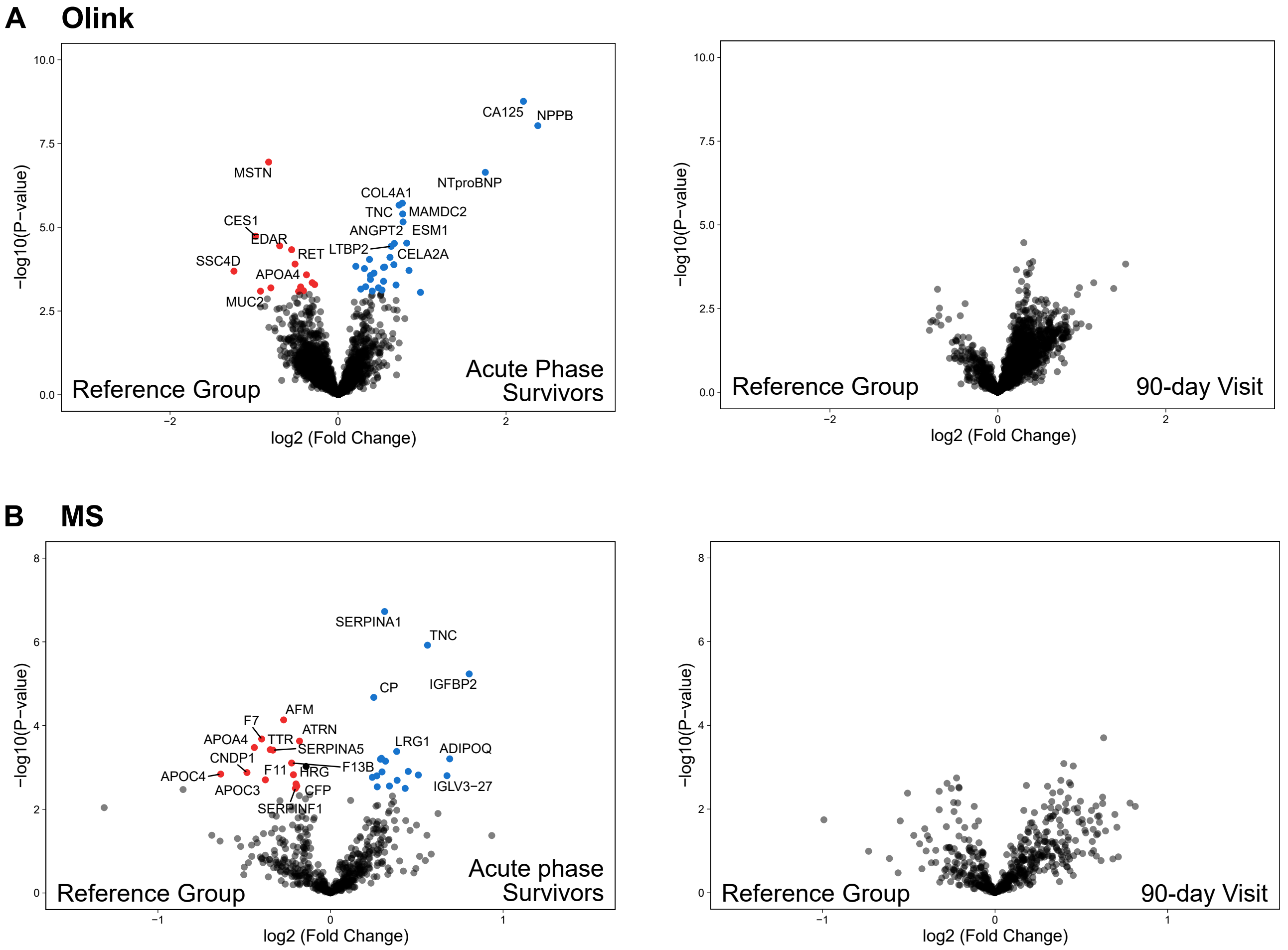
