## Supplementary Table 3 for "Plasma Proteomics in Acute Heart Failure: Survival-Associated Protein Signatures and 90-Day Trajectories"

**Supplementary Table 3**: List of the 20 outcome associated proteins from Cox-regression analysis with the lowest FDR and their potential function in cardiovascular disease.

| Protein | Full name | ln(Hazard Ratio) | 95% Confidence Interval | Adjusted p-Value | C-Index | Role in Cardiovascular Disease | Reference |
|---|---|---|---|---|---|---|---|
| **IGLC2** | Immunoglobulin lambda constant 2 | 1.136 | [0.7, 1.6] | 0.002 | 0.8 | Unknown - Immune Response | |
| **FCRLB** | Fc receptor-like B | 0.783 | [0.4, 1.1] | 0.012 | 0.77 | Unknown - Immune Response | |
| **AFM** | Afamin | -2.241 | [-3.3, -1.2] | 0.013 | 0.81 | Unknown - Decreased in HF patients | Dieplinger et al. 2013 |
| **TMPRSS11D** | Transmembrane protease serine 11D | 1.176 | [0.6, 1.7] | 0.015 | 0.8 | Unknown - Co-expressed with ACE2 | Wruck et al. 2020 |
| **TNFRSF1B** | Tumor necrosis factor receptor superfamily member 1B | 0.956 | [0.5, 1.4] | 0.019 | 0.78 | Inflammation; Has been shown to correlate with poor short-term prognosis in congestive HF. Either provides bioavailable | Dunlay et al. 2008 |

| | | | | | | | |
|---|---|---|---|---|---|---|---|
| | | | | | | TNFalpha or antagonizes its biological activity. | |
| **BAG4** | BAG family molecular chaperone regulator 4 | 1.503 | [0.8, 2.2] | 0.022 | 0.81 | Unknown | |
| **MAD1L1** | Mitotic spindle assembly checkpoint protein MAD1 | 1.502 | [0.7, 2.3] | 0.022 | 0.78 | Unknown | |
| **TXNDC5** | Thioredoxin domain-containing protein 5 | 1.404 | [0.7, 2.1] | 0.022 | 0.79 | Promotes cardiac fibrosis by facilitating ECM protein folding and cardiac fibroblast activation via redox-sensitive JNK signaling. | Shih et al. 2019 |
| **SSBP1** | Single-stranded DNA-binding protein, mitochondrial | 0.665 | [0.3, 1.0] | 0.022 | 0.76 | Potentially reduces cardiac fibroblast proliferation. | Tian et al. 2018 |

| | | | | | | | |
|---|---|---|---|---|---|---|---|
| **INHBC** | Inhibin beta C chain | -1.445 | [-2.2, -0.7] | 0.022 | 0.78 | Unknown - Potentially involved in liver-adipose tissue endocrine axis and thereby causally related to cardiovascular diseases. | Loh et al. 2024 |
| **ZHX2** | Zinc fingers and homeoboxes protein 2 | 1.613 | [0.8, 2.5] | 0.031 | 0.77 | Promotes proinflammatory state in atherosclerosis in mouse model. | Erbilgin et al. 2018 |
| **TNFRSF17** | TNF receptor superfamily member 17 | 1.571 | [0.7, 2.4] | 0.031 | 0.77 | Immune Response - One patient with multiple myeloma recovered from Heart Failure after CAR-T cell therapy against TNFRSF17. | Wang et al. 2024 |
| **ADAMTS4** | A disintegrin and metalloproteinase with thrombospondin motifs 4 | 1.208 | [0.6, 1.8] | 0.031 | 0.78 | Potential biomarker of cardiac injury, positively mediates fibrosis in heart failure models. | Khanam et al. 2022; Vistnes et al. 2023 |

| | | | | | | | |
|---|---|---|---|---|---|---|---|
| **SNRPB2** | U2 small nuclear ribonucleoprotein B" | 0.955 | [0.5, 1.5] | 0.031 | 0.77 | Unknown | |
| **SMNDC1** | Survival of motor neuron-related-splicing factor 30 | 0.678 | [0.3, 1.0] | 0.031 | 0.76 | Unknown | |
| **TNFRSF10A** | Tumor necrosis factor receptor superfamily member 10A | 1.707 | [0.8, 2.6] | 0.033 | 0.78 | Evidence shows association with recurrent acute coronary syndrome. TRAIL signaling is associated with cardiac diseases, and appears protective in atherosclerotic disease. | Vroegindewey et al. 2018; Kelland et al. 2023 |
| **MZB1** | Marginal zone B- and B1-cell-specific protein | 1.091 | [0.5, 1.7] | 0.033 | 0.77 | Not much evidence -May improve mitochondrial function after myocardial infarction. | Zhang et al. 2020 |
| **LBR** | delta(14)-sterol reductase LBR | 1.023 | [0.5, 1,7] | 0.033 | 0.77 | Unknown -Enzyme involved in cholesterol biosynthesis. | Roberti et al. 2003 |
| **BTD** | Biotinidase | -2.605 | [-4.0, -1.2] | 0.033 | 0.79 | Unknown | |

| | | | | | | | |
|---|---|---|---|---|---|---|---|
| **IGFBP2** | Insulin-like growth factor-binding protein 2 | 1.515 | [0.7, 2.4] | 0.034 | 0.77 | Candidate biomarker for HF outcome. | Berry et al. 2015 |
