## Supplementary Table 1 for "Plasma Proteomics in Acute Heart Failure: Survival-Associated Protein Signatures and 90-Day Trajectories"

| **Characteristic** | **Survivors (acute event)**<br>N = 52[1] | **Survivors (90-day follow up)**<br>N = 52[1] | **p-value**[2] |
|---|---|---|---|
| Age (years) | 72 (58, 78) | 72 (58, 78) | |
| Sex | | | |
| Female | 15 (29%) | 15 (29%) | |
| Male | 37 (71%) | 37 (71%) | |
| Clinical Profile | | | |
| Heart Rate (bpm) | 80 (65, 96) | 64 (60, 71) | 0.008 |
| Unknown | 25 | 23 | |
| NYHA | | | |
| NYHA I | 1 (1.9%) | 14 (29%) | |
| NYHA II | 14 (27%) | 22 (46%) | |
| NYHA III | 30 (58%) | 12 (25%) | |
| NYHA IV | 7 (13%) | 0 (0%) | |
| Unknown | 0 | 4 | |
| Laboratory | | | |
| Serum Creatinine (mg/dL) | 1.22 (1.00, 1.50) | 1.20 (0.96, 1.46) | 0.248 |
| eGFR (mL/min/1.73 mp) | 58 (40, 72) | 56 (45, 73) | 0.397 |
| NTproBNP (pg/mL) | 1931 (813, 4,556) | 1040 (472, 2,348) | 0.001 |
| Hemoglobin (g/dL) | 13.30 (11.23, 14.58) | 13.15 (12.08, 14.28) | 0.711 |
| Leukocytes (1000/mm$^3$) | 7.34 (5.72, 9.42) | 7.53 (5.77, 8.73) | 0.619 |
| Thrombocytes (1000/mm$^3$) | 221 (172, 268) | 218 (178, 277) | 0.971 |
| Glucose (mg/dL) | 112 (91, 147) | 110 (94, 129) | 0.512 |
| HbA1c (%) | 6.05 (5.60, 6.50) | 6.10 (5.65, 6.40) | 0.978 |
| Albumin (g/L) | 40.3 (36.6, 42.7) | 43.5 (41.7, 45.6) | <0.001 |
| Total Cholesterol (mmol/L) | 137 (112, 173) | 156 (132, 203) | 0.001 |
| HDL Cholesterol (mmol/L) | 41 (34, 49) | 48 (43, 59) | <0.001 |
| LDL Cholesterol (mmol/L) | 84 (61, 113) | 89 (70, 132) | 0.023 |
| AST (U/L) | 32 (22, 39) | 26 (22, 31) | 0.053 |
| ALT (U/L) | 24 (17, 41) | 21 (14, 29) | 0.039 |
| Gamma GT (U/L) | 72 (39, 132) | 54 (28, 96) | 0.008 |
| Phosphatase Alkaline (U/L) | 80 (69, 99) | 82 (67, 98) | 0.935 |
| LDH (U/L) | 242 (194, 304) | 225 (174, 259) | <0.001 |
| Total Bilirubin (mg/dL) | 0.81 (0.56, 1.07) | 0.55 (0.38, 0.86) | <0.001 |
| Iron (µmol/L) | 13 (8, 15) | 14 (12, 16) | 0.022 |
| Ferritin (µg/L) | 123 (63, 205) | 92 (54, 159) | 0.009 |
| TSH (mU/L) | 2.10 (1.14, 3.50) | 2.01 (1.45, 3.12) | 0.889 |
| Thyroxin (fT4) (pmol/L) | 13.60 (12.41, 15.45) | 13.01 (11.98, 14.40) | 0.397 |
| Sodium (mmol/L) | 142.0 (139.0, 143.0) | 140.0 (138.0, 142.0) | 0.038 |
| Potassium (mmol/L) | 3.80 (3.40, 4.10) | 4.10 (3.90, 4.50) | <0.001 |

| Characteristic | Survivors (acute event) N = 52[1] | Survivors (90-day follow up) N = 52[1] | p-value[2] |
|---|---|---|---|
| Medication | | | |
| ACE/ARB/ARNi | 21 (62%) | 16 (47%) | 0.131 |
| Unknown | 18 | 18 | |
| Betablocker | 33 (97%) | 28 (82%) | 0.074 |
| Unknown | 18 | 18 | |
| Aldosterone Antagonists | 19 (56%) | 14 (41%) | |
| Unknown | 18 | 18 | |
| Loop Diuretics | 21 (62%) | 21 (62%) | |
| Unknown | 18 | 18 | |
| CCB | 7 (21%) | 5 (15%) | 0.617 |
| Unknown | 18 | 18 | |

[1]Median (IQR); n (%)

[2]Wilcoxon signed rank test with continuity correction; McNemar's Chi-squared test; McNemar's Chi-squared test with continuity correction; Wilcoxon signed rank exact test

* At the 90-day follow-up visit, 4 patients in the survivor group had missing laboratory data. Additionally, 3 patients from the survivor group during the acute phase had missing laboratory results.

**Supplementary Table 1.** Patient characteristics comparing the AHF survivors at the acute hospitalization event and at the 90-day follow-up. AHF: Acute Heart Failure.
