## Supplementary Table 2 for "Plasma Proteomics in Acute Heart Failure: Survival-Associated Protein Signatures and 90-Day Trajectories"

**Supplementary Table 2.** Patient characteristics of survivor group with visits at two time points (acute phase and 90-day follow-up) and the reference group

| **Characteristic** | **Reference Group** n = 29 | **Survivors Acute Phase** n = 52 | **Survivors 90-Day Visit** n = 52 |
|---|---|---|---|
| Age (years) | 74 (61, 80) | 72 (58, 78) | 72 (58, 78) |
| Sex | | | |
| Female | 9 (31%) | 15 (29%) | 15 (29%) |
| Male | 20 (69%) | 37 (71%) | 37 (71%) |
| Clinical Profile | | | |
| NYHA* | | | |
| NYHA I | 1 (100%) | 1 (1.9%) | 14 (29%) |
| NYHA II | 0 (0%) | 14 (27%) | 22 (46%) |
| NYHA III | 0 (0%) | 30 (58%) | 12 (25%) |
| NYHA IV | 0 (0%) | 7 (13%) | 0 (0%) |
| Unknown | 28 | 0 | 4 |
| Active/Previous Smoker | 10 (19%) | 26 (50%) | 26 (50%) |
| Unknown | 2 | 0 | 0 |
| Medical History | | | |
| Arterial Hypertension | 23 (79%) | 33 (97%) | 33 (97%) |
| Unknown | 3 | 18 | 18 |
| Diabetes | 29 (100%) | 27 (52%) | 31 (60%) |
| Ischemic Stroke or TIA in the medical history | 3 (10%) | 8 (15%) | 8 (15%) |
| Atrial Fibrillation | 9 (31%) | 34 (65%) | 44 (85%) |
| Peripheral Arterial Disease | 1 (3.4%) | 6 (12%) | 6 (12%) |
| Depression in the medical history | 4 (14%) | 5 (9.6%) | 5 (9.6%) |
| Laboratory* | | | |
| Serum Creatinine (mg/dL) | 1.11 (0.94, 1.53) | 1.22 (1.00, 1.50) | 1.20 (0.96, 1.46) |
| eGFR (mL/min/1.73 $m^2$) | 57 (45, 76) | 58 (40, 72) | 56 (45, 73) |

| | | | |
|---|---|---|---|
| NT-proBNP (pg/mL) | 198 (90, 1235) | 1,931 (813, 4556) | 1040 (472, 2348) |
| Haemoglobin (g/dL) | 13.80 (11.90, 14.80) | 13.30 (11.23, 14.58) | 13.15 (12.08, 14.28) |
| Leukocytes (1000/mm3) | 7.43 (6.36, 8.68) | 7.34 (5.72, 9.42) | 7.53 (5.77, 8.73) |
| Thrombocytes (1000/mm3) | 211 (195, 239) | 221 (172, 268) | 218 (178, 277) |
| Glucose (mg/dL) | 143 (125, 172) | 112 (91, 147) | 110 (94, 129) |
| HbA1c (%) | 7.10 (6.40, 7.60) | 6.05 (5.60, 6.50) | 6.10 (5.65, 6.40) |
| Albumin (g/L) | 42.7 (40.7, 44.3) | 40.3 (36.6, 42.7) | 43.5 (41.7, 45.6) |
| Total Cholesterol (mmol/L) | 138 (119, 168) | 137 (112, 173) | 156 (132, 203) |
| HDL Cholesterol (mmol/L) | 43 (36, 52) | 41 (34, 49) | 48 (43, 59) |
| LDL Cholesterol (mmol/L) | 78 (62, 96) | 84 (61, 113) | 89 (70, 132) |
| AST (U/L) | 25 (20, 33) | 32 (22, 39) | 26 (22, 31) |
| ALT (U/L) | 28 (16, 42) | 24 (17, 41) | 21 (14, 29) |
| GGT (U/L) | 39 (24, 58) | 72 (39, 132) | 54 (28, 96) |
| Phosphatase Alkaline (U/L) | 77 (59, 91) | 80 (69, 99) | 82 (67, 98) |
| LDH (U/L) | 205 (171, 238) | 242 (194, 304) | 225 (174, 259) |
| Total Bilirubin (mg/dL) | 0.60 (0.31, 0.77) | 0.81 (0.56, 1.07) | 0.55 (0.38, 0.86) |
| Iron (µmol/L) | 16 (12, 18) | 13 (8, 15) | 14 (12, 16) |
| Ferritin (µg/L) | 181 (109, 314) | 123 (63, 205) | 92 (54, 159) |
| TSH (mU/L) | 1.61 (0.93, 2.26) | 2.10 (1.14, 3.50) | 2.01 (1.45, 3.12) |
| fT4 (pmol/L) | 13.05 (11.93, 15.71) | 13.60 (12.41, 15.45) | 13.01 (11.98, 14.40) |
| Sodium (mmol/L) | 141.0 (140.0, 143.0) | 142.0 (139.0, 143.0) | 140.0 (138.0, 142.0) |
| Potassium (mmol/L) | 3.85 (3.68, 4.13) | 3.80 (3.40, 4.10) | 4.10 (3.90, 4.50) |

| Medication | | | |
|---|---|---|---|
| ACE/ARB/ARNi | 14 (54%) | 21 (62%) | 16 (31%) |
| Unknown | 3 | 18 | 18 |
| Betablocker | 16 (62%) | 33 (97%) | 28 (82%) |
| Unknown | 3 | 18 | 18 |
| Aldosterone Antagonists | 3 (8%) | 19 (56%) | 14 (41%) |
| Unknown | 3 | 18 | 18 |
| Loop Diuretics | 19 (73%) | 21 (62%) | 21 (62%) |
| Unknown | 3 | 18 | 18 |
| CCB | 8 (31%) | 7 (13%) | 5 (15%) |
| Unknown | 3 | 18 | 18 |

*At the 90-day follow-up visit, 4 patients from the survivor group had missing data for both NYHA (New York Heart Association) class and laboratory results. 3 patients from the survivor group at the acute phase had missing laboratory data only.

*ACE, angiotensin-converting enzyme; ALT, alanine transaminase; ARB, angiotensin receptor blocker; ARNi, angiotensin receptor-neprilysin inhibitor; AST, aspartate transaminase; CCB, calcium channel blocker; eGFR, estimated glomerular filtration rate; fT4, free thyroxine; GGT, gamma-glutamyl transferase; HbA1c, glycated haemoglobin; HDL, high-density lipoprotein; LDH, lactate dehydrogenase; LDL, low-density lipoprotein; NT-proBNP, N-terminal pro-B-type natriuretic peptide; NYHA, New York Heart Association; TIA, transient ischemic attack; TSH, thyroid stimulating hormone.*
